# Mathematical Modeling of Listeriosis Incorporating Effects of Awareness Programs

**DOI:** 10.1101/2020.04.26.20080762

**Authors:** C. W. Chukwu, F. Nyabadza

## Abstract

Awareness programs by the media play a pivotal role in the control of infectious diseases. In this paper, we formulate and analyse a mathematical model for listeriosis incorporating aware individuals. Mathematical analyses of the model are done and equilibrium points determined. The model has three equilibria; namely; the disease-free, the bacteria-free, and the endemic equilibria. Local asymptotic stability of the equilibria is established based on the food contamination number ℛ_*f*_. Numerical simulations are carried out and the effects of various parameters on the model state variables investigated. The results from numerical simulations reveal that an increase in the efficacy of awareness programs, the rate of implementation of awareness programs, and the rate at which unaware susceptible become aware result in the reduction of listeriosis in the human population. The results have important implications in the control and management of listeriosis.

## 1. Introduction

Foodborne diseases (FBDs) are a global public health problem that affects millions of people every year. They are caused by contamination of food products by the various pathogens including bacteria, viruses, and parasites such as *Campylobacter spp*., pathogenic *Escherichia coli, Salmonella spp*., *Vibrio, Yersinia* and *Listeria monocytogenes* (*L. monocytogenes*) [1]. Foodborne listeriosis is caused by the bacteria Gram-positive bacillus *L. monocytogenes*. This organism can be found in the environment such as soil, water or vegetation e.t.c. In particular, the primary source of human *Listeria* infection is either from the consumption of contaminated ready-to-eat food products or directly from the environment [2]. This bacteria is successful in causing listeriosis because it survives food processing technologies that rely on acidic or salty conditions, unlike many other pathogens. It continues to multiply slowly at low temperature which allows its growth even in properly refrigerated food products[3, 4]. It is a relatively rare disease with 0.1 to 10 cases per 1 million people per year depending on the countries or regions of the world. For example, the case fatality ratio in South Africa 2017/2018 outbreak is estimated at 28.6% compared to worldwide listeriosis outbreaks. Even though the number of cases of listeriosis is relatively small, the high death rate and management costs associated with this disease make it a significant public health concern [5].

Recently, many mathematical models incorporating awareness programs by media have been formulated and analysed, to predict the impact of awareness programs in preventing or controlling infectious diseases [8, 20, 9, 10, 11, 12, 13, 14, 15, 21, 28, 23]. In particular, Misra et al. [31] assumed that during the spread of an infectious disease, aware individuals avoid contacts with the infectives by taking precautionary measures during the infection period. In [14], a model with the intensity of media campaign proportional to the number of the infected individuals was assumed and analysed. Information regarding the protection against the disease is propagated through television (TV), and social media advertisements (SMA‘s). The growth rate of the cumulative number of TV and SMA‘s was assumed to be proportional to the number of infected individuals and aware individuals. However, the growth rate against the aware individuals, (*A*), decreases with a factor 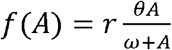. The study suggests that the net growth rate of TV and SMA‘s too aware population becomes 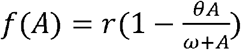, where *θ* is a constant denoting the decay in advertisements due to increase in the number of aware individuals and *ω* represents the half-saturation point for this interaction as f(A) attains half of its maximum value *rθ* due to the effect of TV and SMA‘s [28]. Their results show that the increment in the growth rate of TV, and SMA‘s destabilizes the system, and periodic oscillations arise through Hopf-bifurcation. Also, TV and SMA‘s during the spread of infectious diseases have the potential to bring behavioural changes among the people and control the spread of the diseases. Imbusi et al. [23], studied the effect of media campaigns on Jiger infestation. They assumed that the aware individuals interact with the infected individuals and hence get infected. Bifurcation analysis of their study reveals that the model has an intrinsic backward bifurcation whenever the parameter that accounts for the proportion of larvae which develop into adult female fleas is involved in jiggers transmission. Furthermore, an increase in media campaign decrease rate of contracting Jiggers infections. On the other hand, mathematical models incorporating awareness programs on disease transmission rates were analysed in [16, 20, 17]. Authors in [17], used the transmission rate with media effect represented by 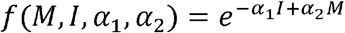, where *M* denotes the media campaigns, *I* the infective individuals, *α*_1_, and *α*_2_ are the weights of media effect sensitive to infectives, and media campaigns respectively. Results from simulation reveal that awareness programs reduce the number of cases during an epidemic, though the number of cases is further mitigated under some optimal reporting intensity. Sensitivity analysis of their model also reveals that an outbreak severity is more sensitive to the weight *α*_1_ than weight *α*_2_.

Further, a limited number of mathematical models on listeriosis disease transmissions dynamics have been done, see for instance [30, 24, 26, 25, 27]. However, to the authors best knowledge, none of these mathematical models has considered the effects of awareness programs on the control of listeriosis to date. In the present study, we formulate a mathematical model for listeriosis infection from consumption of contaminated food products incorporating awareness programs and assuming that the aware individuals interact with the infectives as in [23]. This paper aims to study the effects of imperfect awareness programs using a deterministic listeriosis epidemic model.

This manuscript is organized as follows: In Section 2, a mathematical model that describes the effect of the imperfect awareness program on listeriosis epidemic is formulated and its basic properties in Section 3. In Section 4, the model equilibria were found and their local stability discussed. Numerical simulations confirming some of the analytical results were presented in Section 5. Finally, a brief discussion and conclusion are given in Section 6.

## 2 Mathematical model

In this section, a deterministic model that describes the effect of imperfect awareness programs on listeriosis epidemic. The model consists of four sub-components, namely; the human population, bacteria population, awareness program, and the food products. Let *A*_*p*_(*t*) represent the cumulative density of awareness program driven by media at any time t, A the maximum intensity of *A*_*p*_(*t*), with *µ* the rate of implementation, and *µ*_0_ the rate of diminution of awareness program over time. The human population is divided into four sub-classes namely; unaware susceptible, *S*_*u*_(*t*), (individuals who are prone to contracting listeriosis), aware susceptible, *S*_*a*_(*t*), (individuals who are become aware during disease spread as a result of awareness programs), infected, *I*(*t*), (individuals who are infected with listeriosis disease), recovered *R*(*t*), (individuals who would have recovered from listeriosis through treatment). The total human population *N*(*t*) at any time *t* is thus given by

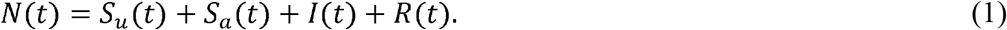

The unaware individuals are recruited into the human population at a rate proportional to the human population so that we have *µ*_*h*_*N* unaware susceptible individuals become infected at a rate 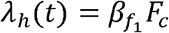, and move to the infectious class who then recover at a rate *α* with no immunity and become susceptible to the disease again [6]. Also, the susceptible individuals move into the aware class at a rate *ω*_l_ as a result of the awareness programs, whom in-turn become unaware susceptibles again at a rate *ω*_2_ due to waning of the awareness programs. We assume the awareness programs are not 100% effective so that the awareness susceptibles are infected at a rate (1 − *ε*) *λ*_*h*_(*t*), where the efficacy parameter, 0 ≤ *ε* ≤ 1, accounts for the effectiveness of the media programs in protecting the aware individuals. If *ε* = 1, then the awareness programs are not effective, and *ε* = 0 corresponds to completely ineffective awareness programs, while 0 < *ε* < 1 means that the awareness programs will be effective to some extent. In this model, we assumed that the recovered individuals do not contribute to disease transmissions, and the growth rate of the cumulative density of awareness program is proportional to the number of infected individuals. Also, we assume a natural mortality rate *µ*_*h*_ for all the human compartments.

We define *B*(*t*) to be the bacteria population in the environment with a carrying capacity *K*_*B*_, and a net growth rate *r*_*b*_. On the other hand, we define the uncontaminated food products by, *F*_*u*_(*t*), and the contaminated food products by, *F*_*c*_ (*t*), so that the total amount of food products at any time *t* is given by

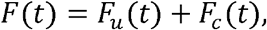

with the addition and the rate of removal of food products *µ*_*f*_. Uncontaminated food *F*_*u*_(*t*) are contaminated by the bacteria from the environment at a rate 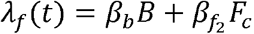, and they become contaminated. These contaminated food products then cause human listeriosis which is as a result of ingestion of contaminated foods by humans at a rate denoted by *λ*_*h*_(*t*) = *β*_l_*F*_*c*_. Combining the model; parameters, assumptions, and description above, we have the following nonlinear system of ordinary differential equations:

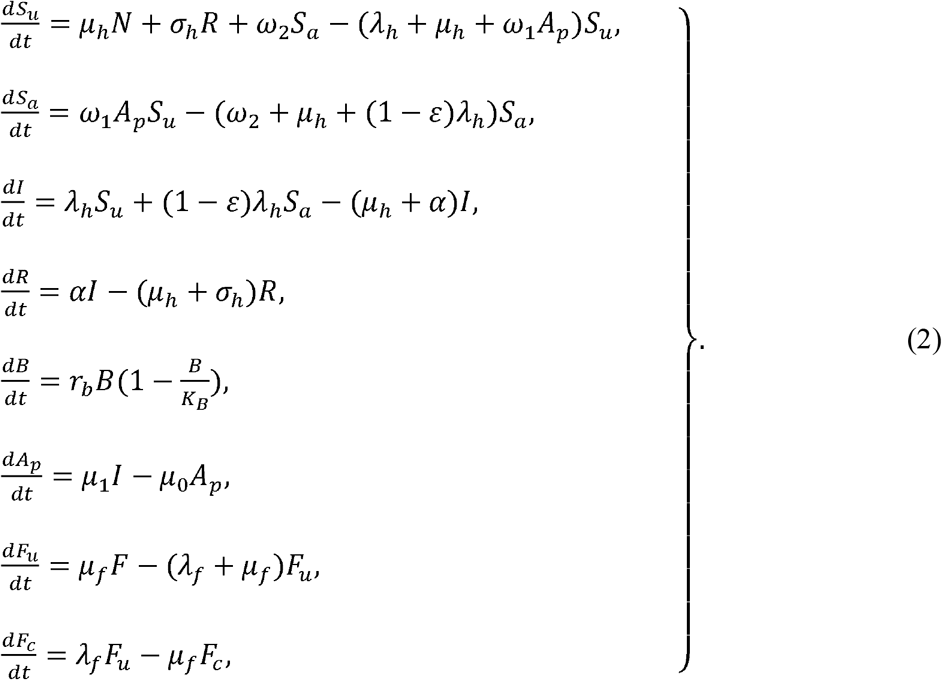

We substitute *R* (*t*) = *N*(*t*) − *S*_*u*_ (*t*) − *S*_*a*_ (*t*) − *I*(*t*) from (1), and non-dimensionalise system (2) by setting

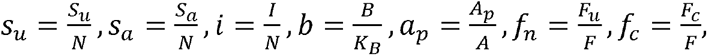

and obtain the following system of equation

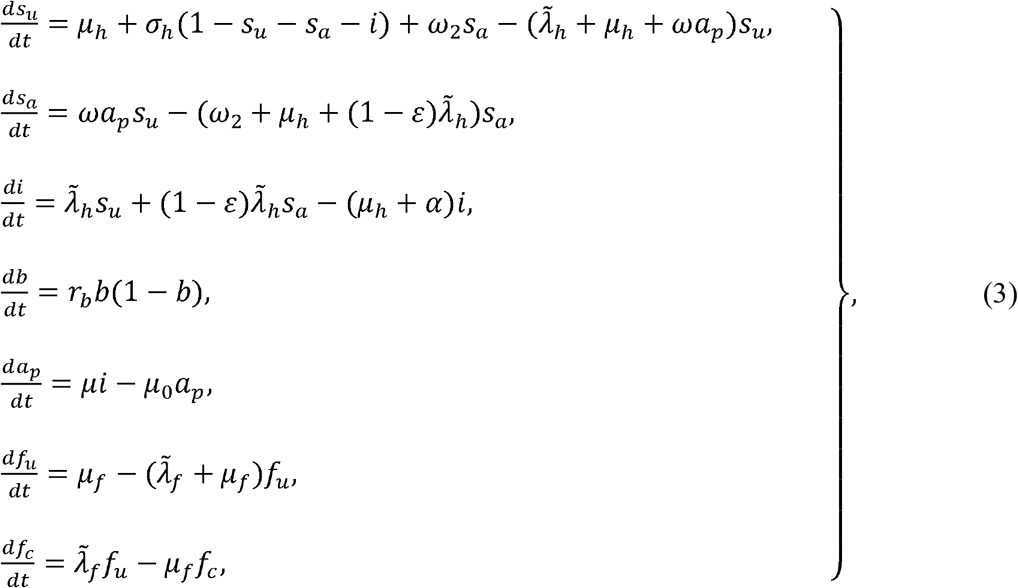

where

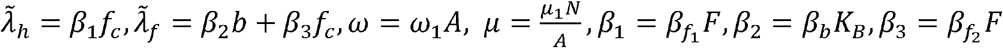

subject to following initial conditions

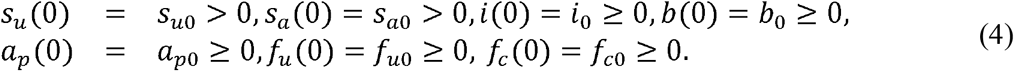

The model parameters for system (3), are assumed to be non-negative for all time *t* except for the net growth rate *r*_*b*_.

## 3 Model analysis

### 3.1 Existence of solutions

Let Γ be the epidemiological biological meaningful region for system (3) contained in 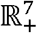. Firstly, we prove that the solutions exist and are bounded. We, therefore, have the following theorem on the existence of solutions.

#### Theorem 1

*The solutions of model system (3) are contained in the region* 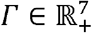, *which is given by* 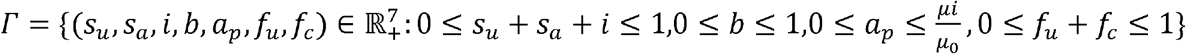, *for the initial conditions (4) in* Γ.

*Proof*. The total change in human population from the model equations (3) is given by

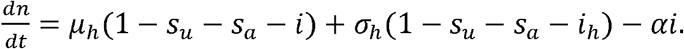

Since *n*= *s*_*u*_ +*s*_*a*_ +*i* ≤ 1, we set *ψ*= (*s*_*u*_ +*s*_*a*_ +*i*_*h*_) and obtain

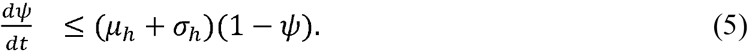

Solving the differential inequality (5), yields

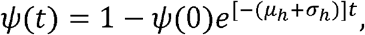

with *ψ* (*t*) ≤ 1 as *t* → ∞. Thus, the upper bound of *ψ* (*t*) is 1 provided that *ψ* (0) 2 ≥ 0. Consider

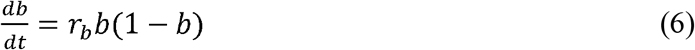

the bacteria population is given by the fourth equation of model system (3). The solution of (6) yields 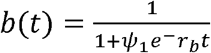, with *b*(*t*) ≤ 1 as *t* → ∞. Thus, *b*(*t*) is bounded above by 1. We emphasize that the bacteria grows logistically, and since it exists naturally in the environment it can easily cause contamination of the ready-to-eat food products which leads to human listeriosis.

On the other hand, the total change in the number of food products is given by summing the second and last equation of model equation (3) which is

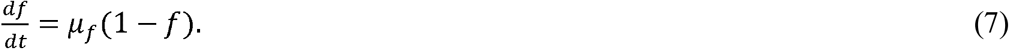

The solutions of (7) yields

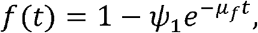

where *ψ*_1_ is a constant with *f*(*t*) ≤ 1 as *t* → ∞, also *f*(*t*) is bounded above by 1. Hence, all the solutions of model system (3) exists, are epidemiological meaningful, bounded, and remain in Γ for *t* > 0.

### 3.2 Positivity of solutions

We show that the solutions of system (3) are positive subject to the following initial conditions (*s*_*u*_(0), *s*_*a*_(0), *I*(0), *b*(0), *a*_*p*_(0), *f*_*u*_(0), *f*_*c*_ (0))^*T*^ ∈ ℝ^+7^. We use Lemma 1 as in [7], stated as follows.

#### Lemma 1

*Suppose Γ* ⊂ ℝ × *C*^*n*^ *is open, f*_*i*_ ∈ *C*(*Γ*, ℝ), *i* = 1, …, *n. If*

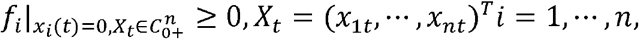

then 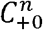 is the invariant domain of the following equations

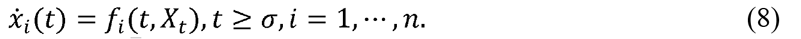

If 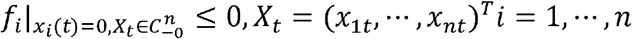 then 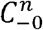 is the invariant domain of equations (8). We have the following result on the positivity of solutions of model system (3).

#### Theorem 2

*The solutions* (*s*_*u*_ *(t), s*_*a*_ *(t),I (t),b (t),a*_*p*_ *(t),f*_*u*_ *(t) f*_*c*_ *(t)*) *of the model system (3) with positive initial values (4) are positive for all t* ∈ (0, ∞)

Let *X* = (*s*_*u*_,*s*_*a*_,*I,b,a*_*p*_,*f*_*u*_,*f*_*c*_) ^*T*^ and

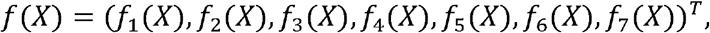

then we can re-write the model system (3) as follows

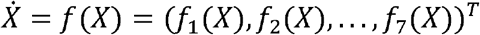

where

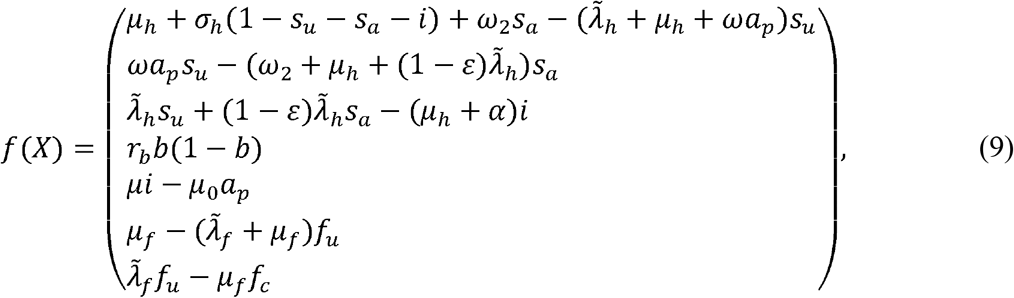

with (•)^*T*^ being the transpose. Setting all the model state variables in (9) to zero gives

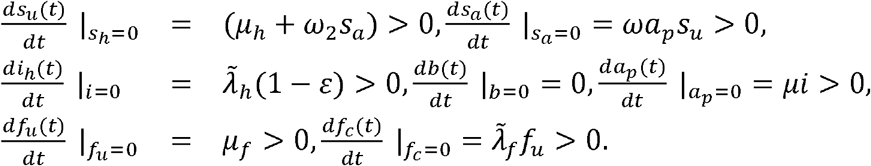

Hence, by Lemma 1 as in [7], we thus have that 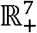 is positively invariant set.

## 4 Equilibria and their stability analysis

### 4.1 Equilibria

We set the right side of equations (3) to zero as follows

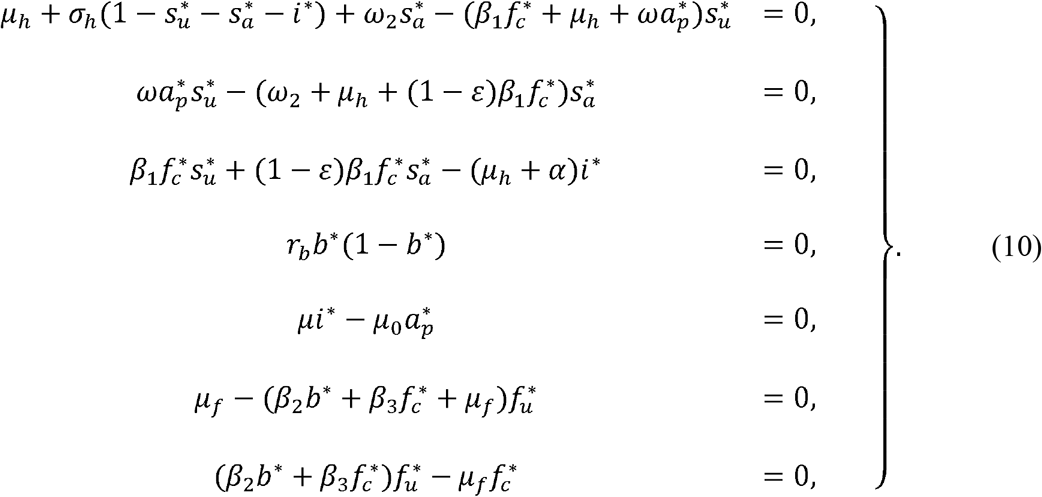

Solving system (10) for the equilibria, from the fourth equation of system (10) we have that

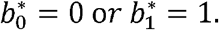

Firstly, we consider the case when *b*\* = 0.

**Case I:** If 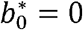,the second last equation of (10) gives that

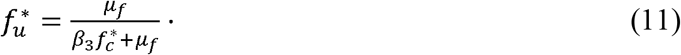

Substituting (11) into the last equation of (10) we obtain

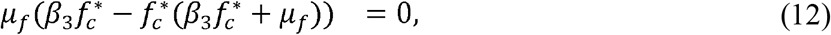

and after some algebraic manipulation, we obtain

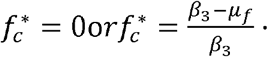

Hence, when 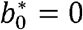 we have that 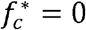, and as a result 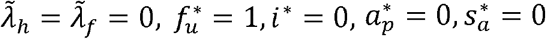 and 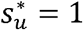.

This results in the disease-free equilibria (DFE) denoted by

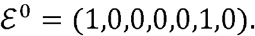

Further simplification of the nonzero 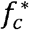 yields

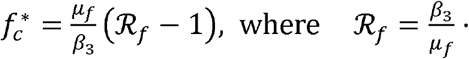

Thus, we have the following result on the existence of the non-zero equilibrium point 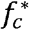.

#### Lemma 2

*The nonzero equilibrium point* 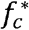 *exists if and only if ℛ*_*f*_> 1.

Here, we emphasize that ℛ_*f*_ denotes the *food contamination threshold* from the contaminated food to the uncontaminated food products. This is equivalent to the basic reproduction number *ℛ*_0_, in disease modelling as defined in [32, 33]. However, we note that 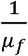 is the duration of stay in the food compartments. Thus, ℛ_*f*_ represents the average amount of food products that can be contaminated and responsible for causing human listeriosis.

When 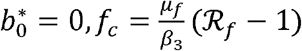, from the third and fifth equations of (10) we have

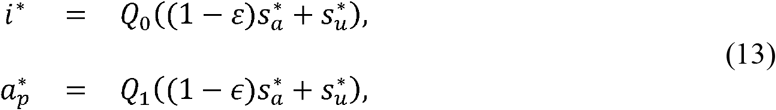

where 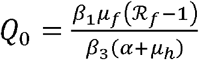 and 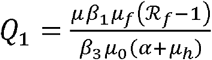 Substituting (13) into the second equation of system (10), and express 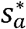 in terms of 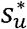, we have

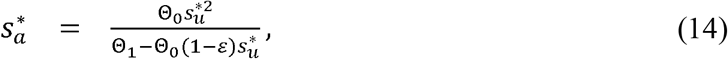

where Θ_0_ = *μωβ*_1_*μ*_*f*_ (ℛ_*f*_ − 1) and Θ_1_ = *μ*_0_(*α+ μ*_*h*_) (*β*_1_*μ*_*f*_ (1 − *ϵ*) (ℛ_*f*_ − 1) + *β*_3_ (*μ*_*h*_ + *ω*_2_)) Substituting (14) into the first equation of (10) gives the quadratic equations

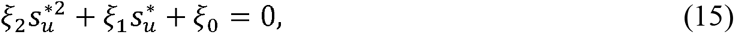

where

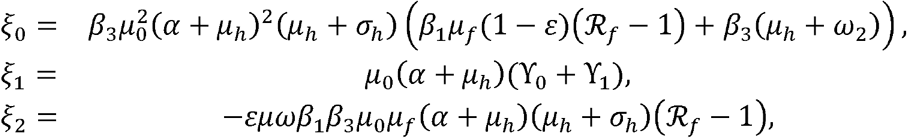

with

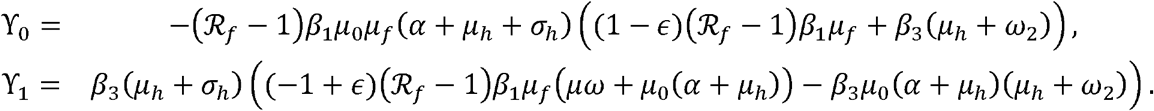

Note that, *ξ*_0_ > 0, *ξ*_2_ < 0 if ℛ_*f*_ >1. Applying Descartes’ rule of sign change, equation (15) has only one possible positive solutions for 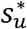 irrespective of *ξ*_1_ the sign of Thus, we have the bacteria-free equilibrium point 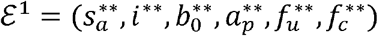,

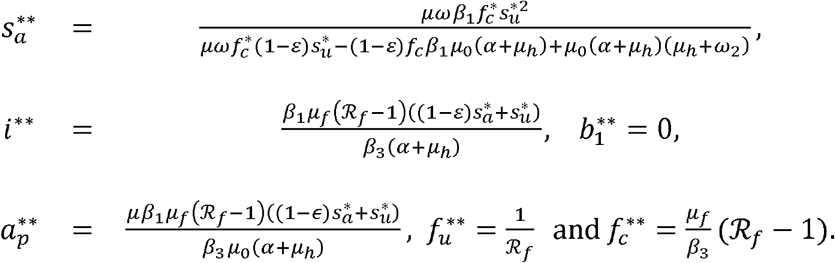

Before considering the second case when *b*\* =1, we first look at the local stabilities of the DFE and the LFE points.

#### 4.1.1 Local stability of the disease-free equilibrium point

We state the following theorem for the local stability of *ε*^0^.

##### Theorem 3

*The equilibrium point ε*^0^ *is locally asymptotically stable if r*_*b*_ < 0 *and ℛ*_*f*_ < 1 *and unstable otherwise*.

*Proof*. We show that our system is locally asymptotically stable by evaluating the Jacobian matrix at *ε*^0^ as follows

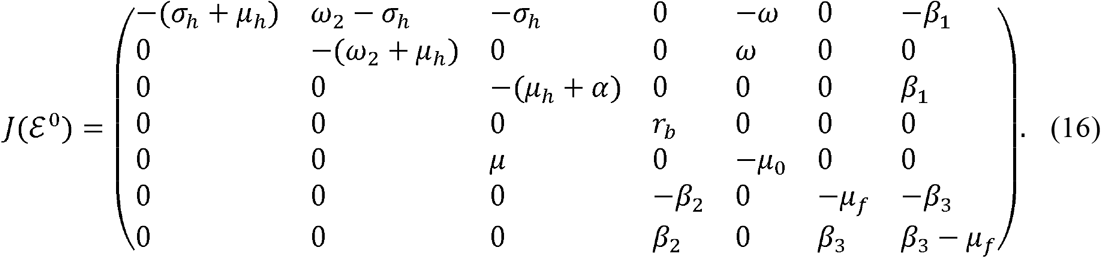

Thus, the eigenvalues of *J*(*ε*^0^) are; − (*σ*_*h*_ + *μ* _*h*_), − (*ω*_2_ + *μ* _*h*_), −(*μ*_*h*_ + *α*),*r*_*b*_, − *μ*_0_ The remaining eigenvalues are determined from the determinant of

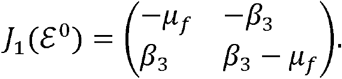

and are obtained from the solutions of the quadratic equation

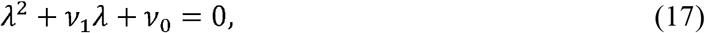

with

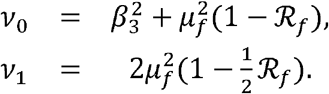

If ℛ_*f*_ < 1,then *v*_1_ > 0, and *v*_0_ > 0. Hence, the eigenvalues of (17) have negative real parts by the Routh-Hurwitz stability criterion. We note that if *r*_*b*_ < 0, all the eigenvalues at *ε*^0^ are negative, hence *ε* ^0^ is locally asymptotically stable. However, if *r*_*b*_ > 0, then *ε*^0^ is unstable. Note that if *r*_*b*_ < 0 then Listeria in the environment dies out, resulting in a locally asymptotically stable equilibrium point, while it establishes itself when *r*_*b*_ positive resulting in an unstable equilibrium point.

#### 4.1.2 Local stability of the bacteria-free equilibrium point

We state the following theorem for the local stability of *ε* ^1^.

##### Theorem 4

*The equilibrium point ε*^1^ *is locally asymptotically stable if r*_*b*_ < 0 *and ℛ*_*f*_ >1, *and unstable otherwise*.

*Proof*. We show that our system is locally asymptotically stable by evaluating the Jacobian matrix at *ε*^1^ as follows

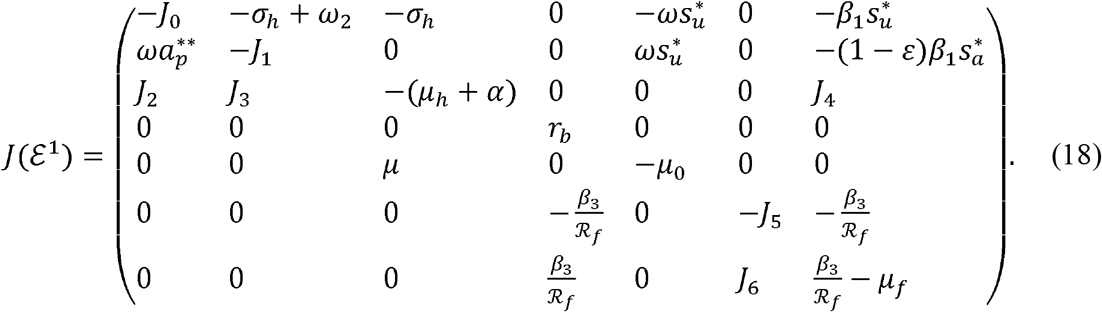

for positive constants *J*_*k*_′s, *k* = 0 …,6, if the value for ℛ_*f*_ > 1 and given bye

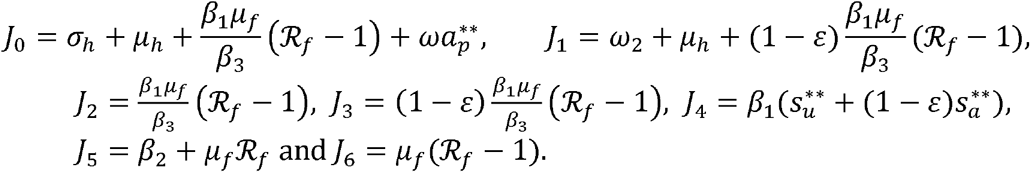

The eigenvalues of from matrix (18) are; *r*_*b*_, − *μ*_0_ and those from matrices

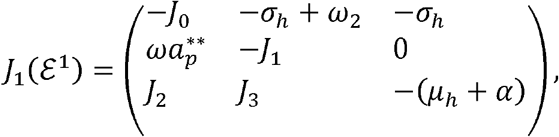

and

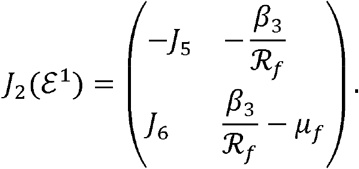

The eigenvalues from 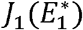 are given by the cubic equation

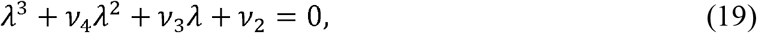

for

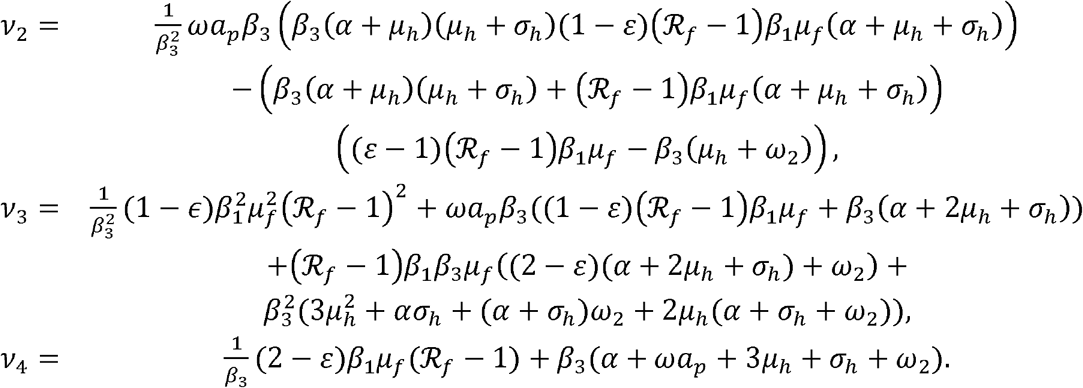

Given that, all the coefficients of *v*_2_, *v*_3_, *v*_4_ are positives, and

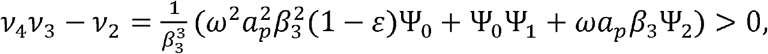

for

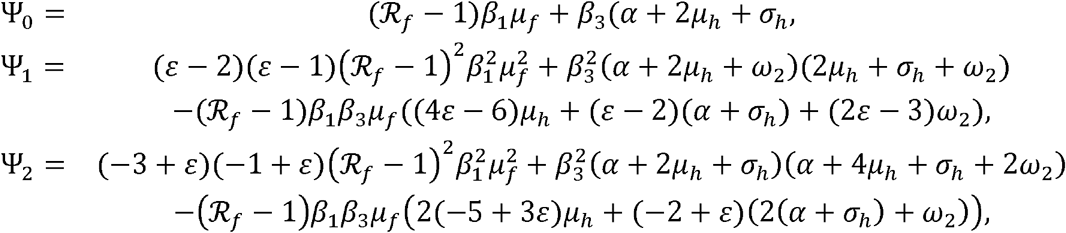

Thus, the roots of equation (19) have negative real parts by the principle of Routh-Hurwitz stability criterion. Next, we consider the matrix 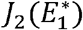. Its eigenvalues are obtained from the quadratic expression

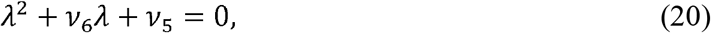

given that

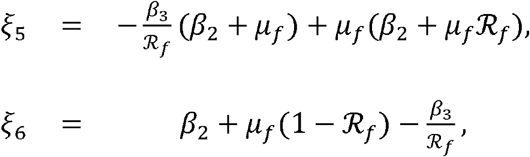

Note that, provided the value of ℛ_*f*_ < 1, we will have that *ξ*_5_, and *ξ*_6_ are positives. Thus, making the eigenvalues of *J*_2_ (*ε*_1_) to have negative real parts. However, *J*(*ε*_1_) will be locally asymptotically stable if and only if the eigenvalue *r*_*b*_ < 0, and unstable if *r*_*b*_ > 0.

**Case II:** Secondly, when 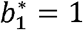 solving for 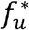 from the sixth equation of (10), we have that

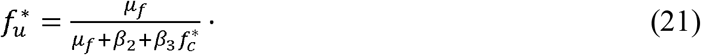

Substituting (21) into the last equation of (10) we obtain the following quadratic expression after some algebraic simplifications

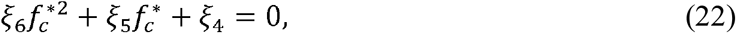

where

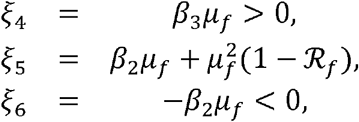

We note that *ξ*_5_ >0 if ℛ_*f*_ <1 or *ξ*_5_ < 0 ℛ_*f*_ > 1. Thus, irrespective of the signs of *ξ*_5,_ the quadratic equation (22) has one positive solution for 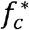 say 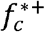. Similarly, from the third, and fifth equations of (10) we have

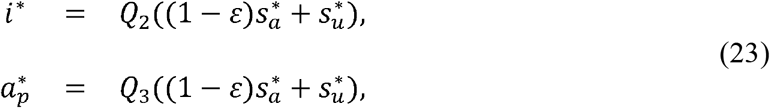

where 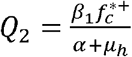 and 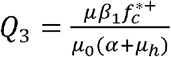 Substituting (23) into the second equation of (10) and expressing 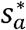 in terms of 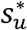, we obtain the following expression after some algebraic manipulations

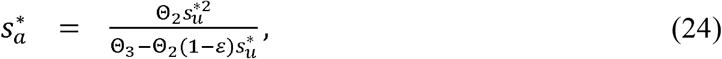

where 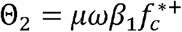 and 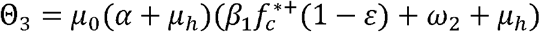. Substituting (24) into the first equation of (10), we obtain the following quadratic equation

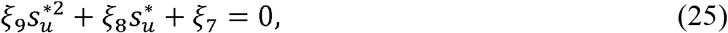

where

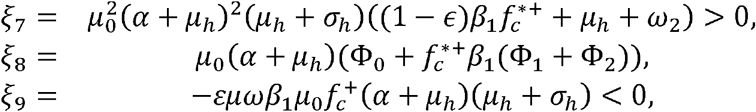

with

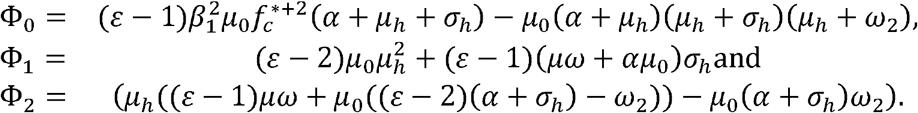

Note that, Φ_0_ < 0, Φ_1_ < 0, and Φ_2_ < 0 making *ξ*_8_< 0. Hence, the solution to equation (25) given by

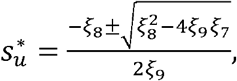

have one positive solution for 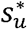. We thus have the following result on the existence of new 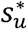 called 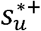.

##### Lemma 3

*The equilibrium point* 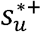 *exists subject to the existence of* 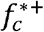.

Thus, we have the endemic equilibria 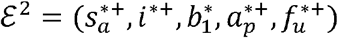 given by

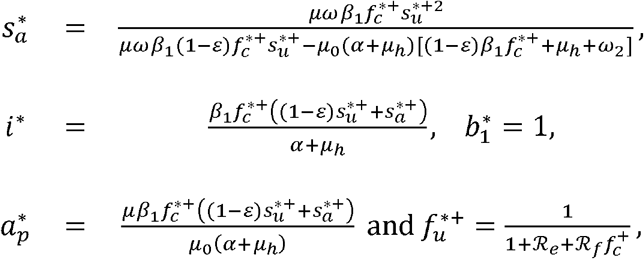

where

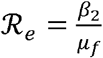

denotes the contamination threshold contributed by the environment.

#### 4.1.3 Local stability of the endemic equilibrium point

Theorem 5 governs the existence of the local stability of the endemic equilibria.

##### Theorem 5

*The equilibrium point ε* ^2^ *is locally asymptotically stable if ℛ*_*f*_ > 1, *r*_*b*_ > *o and unstable otherwise*.

*Proof*. Evaluating the Jacobian at *ε*^2^, we have that

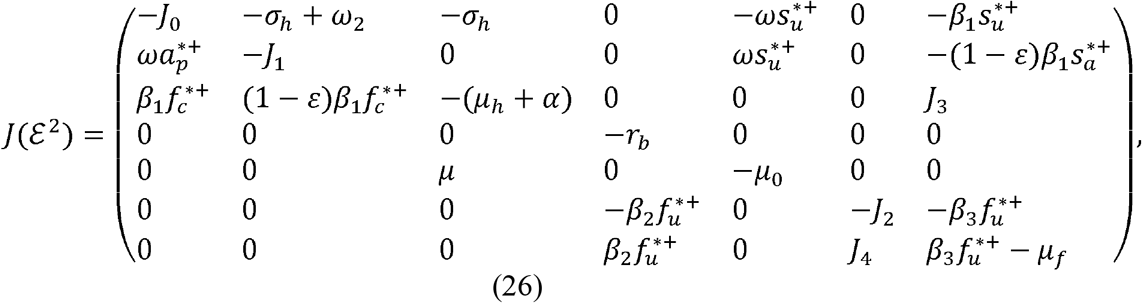

where the *J* ′_*j*_ s, *j* = 0, …, 4, are positive and given by

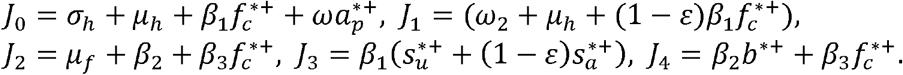

It can be seen that the eigenvalues from (26) are; − *r*_*b*_ and − *μ*_0_ The rest eigenvalues are determined from two matrices

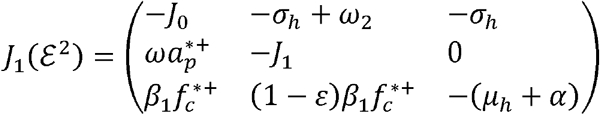

and

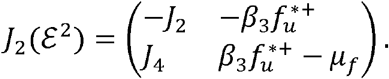

The eigenvalues of 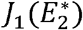 are obtained from the cubic equation

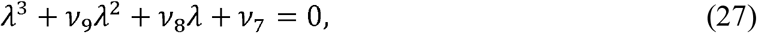

where

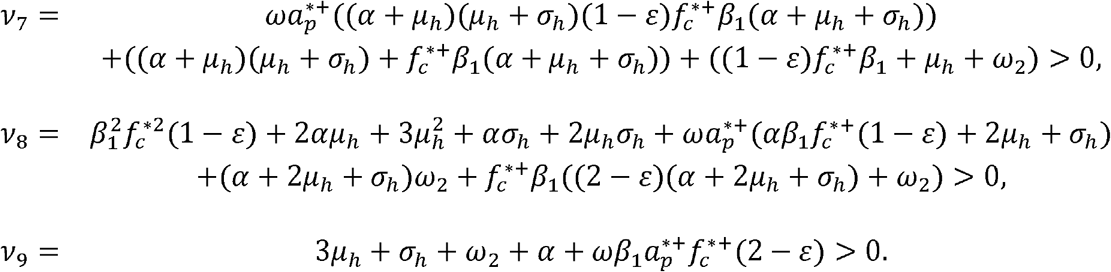

Given all the coefficients are positive and

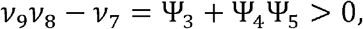

where

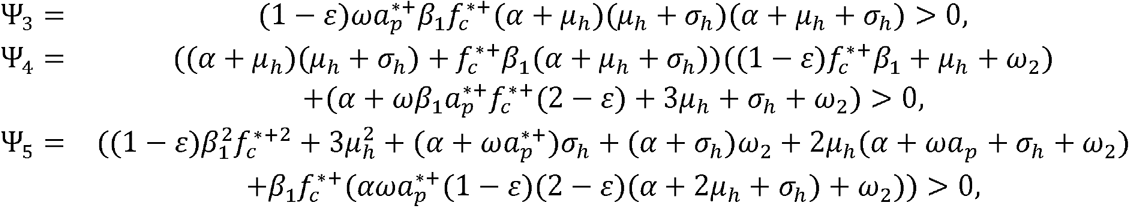

Hence, the roots (27) have negative real parts by the Routh-Hurwitz criterion. We now focus our attention on *J*_2_ (*ε*^2^) The remaining eigenvalues are obtained from the quadratic equation

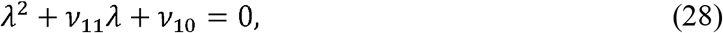

where

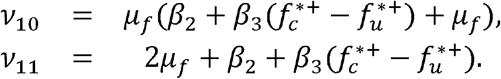

Note that *v*_10_ > 0, *v*_11_ > 0 if 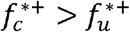. Hence, by Routh-Hurwitz criterion the eigenvalues of (28) have negative real parts. Given that *r*_*b*_ > 0, *ε*^3^ is locally asymptotically stable. However, if *r*_*b*_ < 0 then *ε*^3^ is unstable. The biological significance of 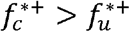, implies that, so long there are more contaminated food products *f*_*c*_ (*t*) compared to uncontaminated food products *f*_*u*_(*t*), listeriosis infection will always reach its endemic state in the human population. Thus, making it difficult for the disease to be contained.

## 5 Numerical simulations

In this section, we present numerical simulations of the model system (3) confirming most of the analytical results obtained in this manuscript. We begin by looking at the estimation of the parameters for the model. There are very few mathematical models on human listeriosis disease, as a result, it difficult to find the model parameters in literature and research articles. Thus, most of the parameters’ values were estimated for the work done in this paper except for the diminution rate of the awareness campaign, *µ*_0_, and the human natural mortality rate *µ*_*h*_, which are taken from [29] and [30] respectively. The sets of parameters that give rise to the endemic equilibrium points are given in Table 1. The simulations of model system (3) were carried out using fourth-order Runge-Kutta numerical method scheme implemented in Matlab with a unit time step 1 to perform the simulations with the following initial conditions for the state variables;

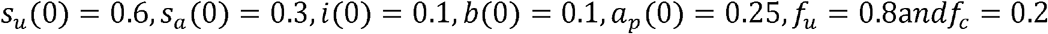

also reflected at the endemic equilibria. These numerical values were hypothetically chosen, and do not represent any real-life scenarios and are thus for illustrative purposes only.

**Table 1:**
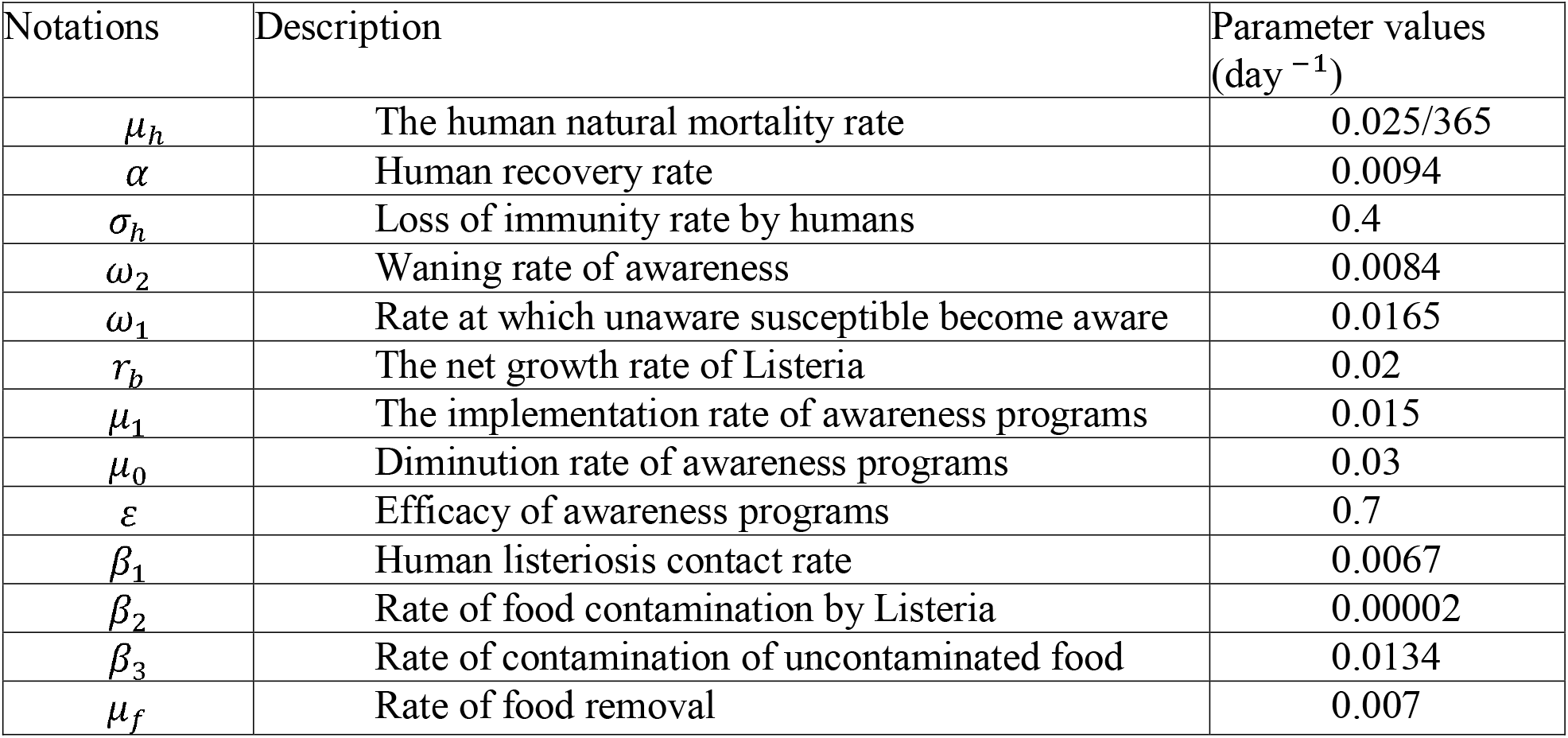
Description and estimation of parameter values used in the simulations of model system (10).

### 5.1 Convergence of equilibrium points

In Figure 1 we present a phase portrait confirming the analytical results for the local stability of the DFE and the EE points respectively. Figure 1(a) shows the relationship between *f*_*c*_ and *i* as their initial conditions are varied. We observe that the trajectories tend to the point 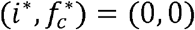 irrespective of the starting point. Also, Figure 1(b) shows the phase portrait of *a*_*p*_ against *i* for different initial conditions with all the trajectories tending to the endemic equilibrium 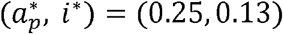, with ℛ_*f*_ = 1.9143.

**Figure 1:**
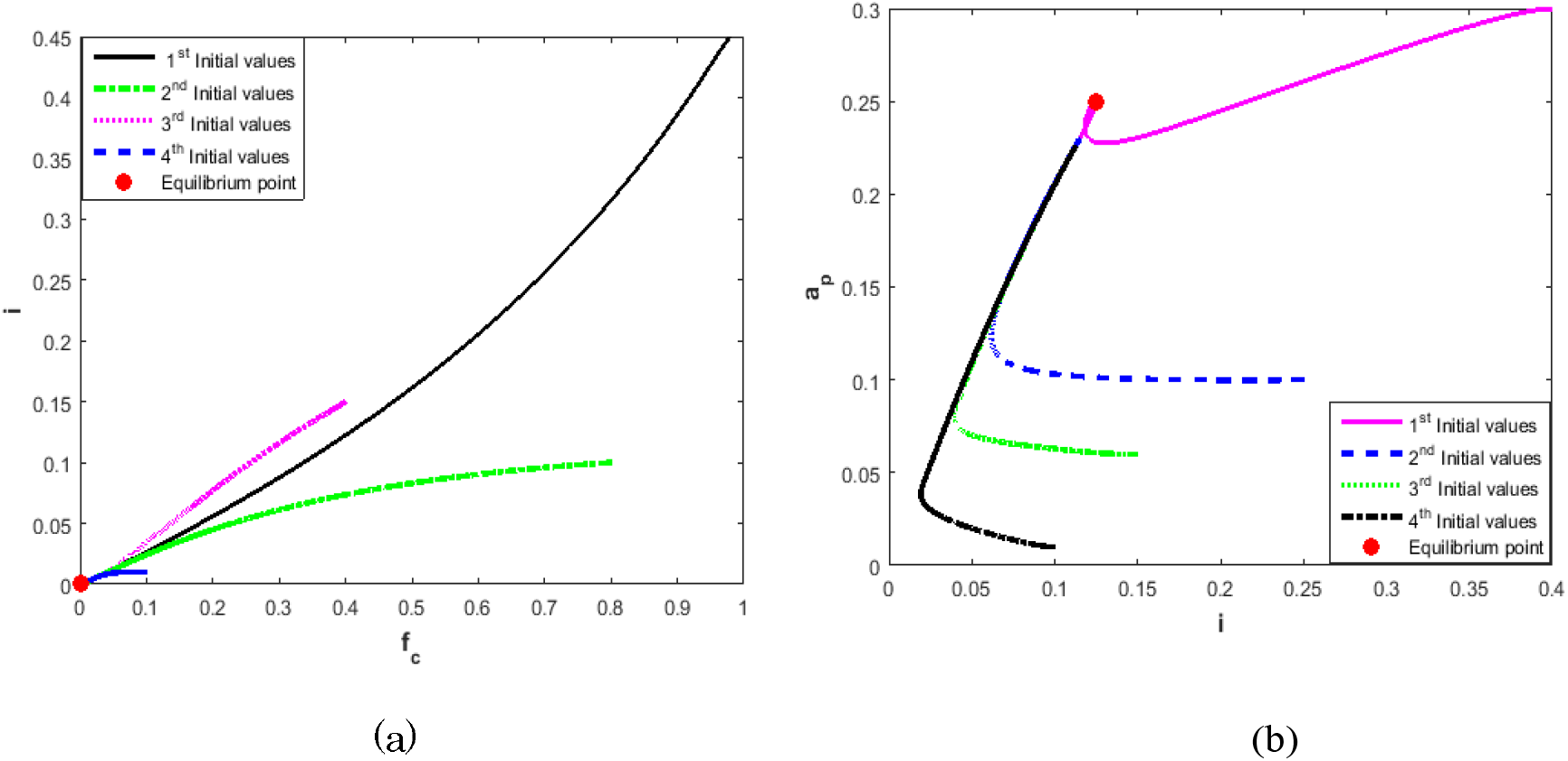
(a) Phase portraits of the disease-free equilibria points for *is*_*a*_ -plane with varying initial conditions, and corresponds to ℛ_*f*_ = 0.5333. (b) Phase portraits of the bacteria-free equilibria points for *is*_*u*_-plane with varying initial conditions, and corresponds to ℛ_*f*_ = 1.5714. (c) Phase portraits of the endemic equilibria points for *ia*_*p*_-plane with varying initial conditions, and corresponds to ℛ_*f*_ = 1.9143.

### 5.2 Sensitivity and uncertainty analysis

Sensitivity analysis is used to measure the relative change in a state variable when a given parameter changes. Latin-hypercube sampling method of stratified uncertainty analysis [34], was used to determine model parameters that are sensitive model system (3). The listeriosis model parameters were simulated against the state variable *i*. The simulation was done using Matlab with 1000 runs over 1200 days, and its Tornado and scatter plots for parameters with positive, and negative partial rank correlation coefficients (PRCC‘s) shown in Figures 2 and 3 respectively. The implications of increasing model parameters; *ω*_2_, *µ*_0_ with negative PRCC‘s will negatively impact the infected individuals. On the other hand, an increase in model parameters such as; *µ, ω* and *ϵ* will positively impact human listeriosis disease cases. Furthermore, an increase in *ω*_2_ will lead to more individuals losing awareness, resulting in an increased risk of been exposed and getting infected due to waning.

**Figure 2:**
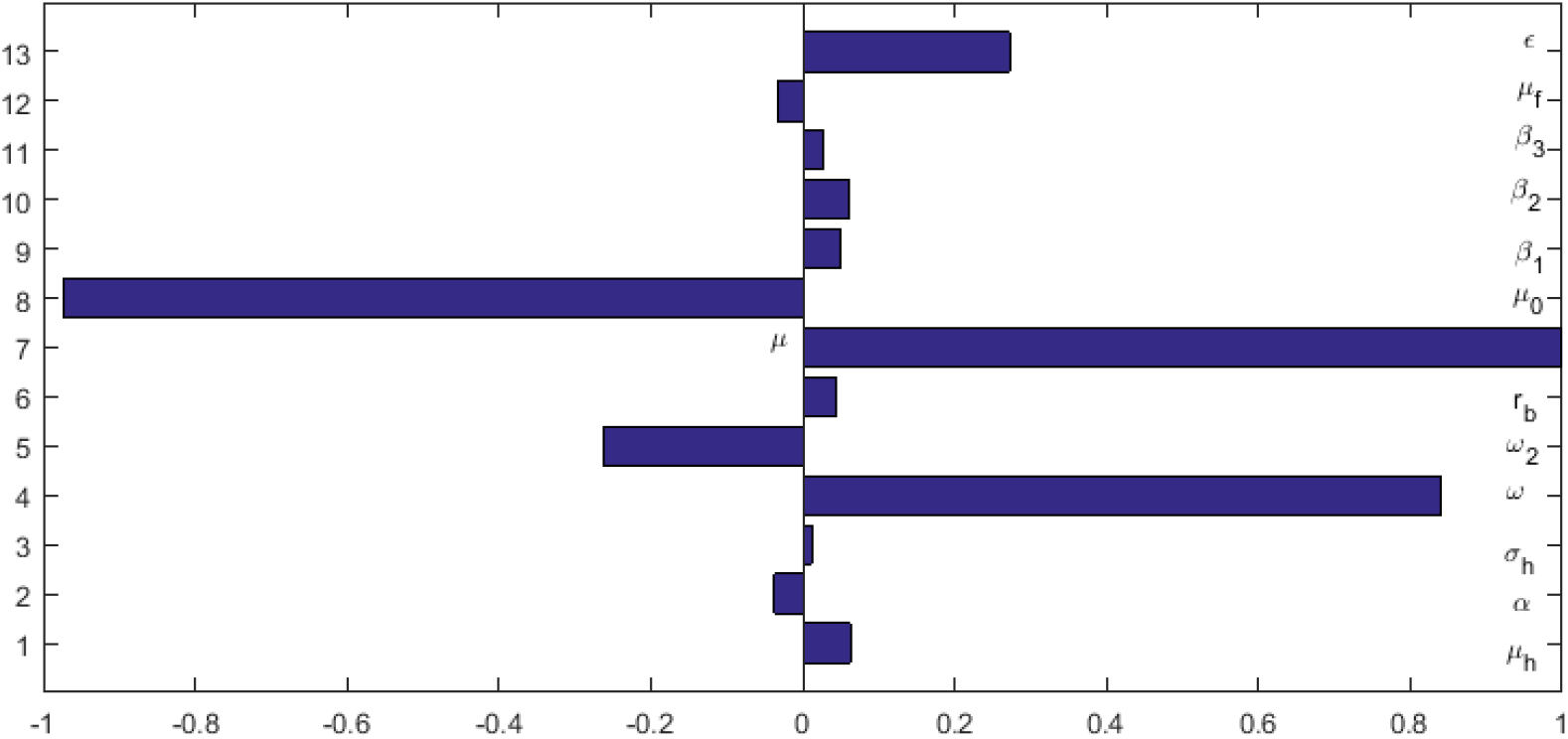
Tornado plot of all model parameters driving listeriosis disease dynamics. Note that *ε* is represented by *ϵ* in the legend.

**Figure 3:**
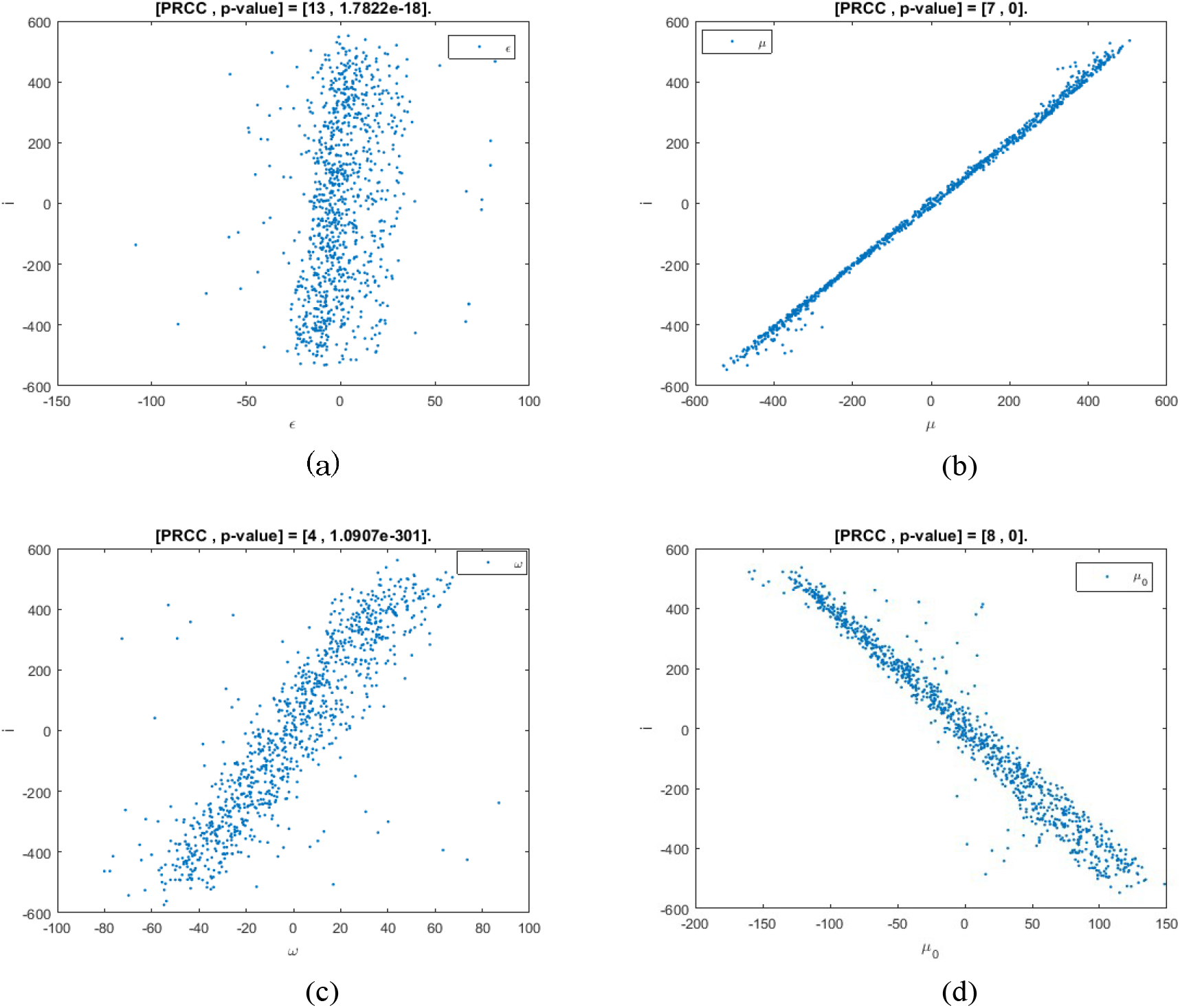
(a), (b), (c) scatter plots of; parameters *ε, ω*_2_, *µ* with positive partial correlation coefficients versus i -individuals while Figure (d) the parameter *µ*_0_ have negative partial correlation coefficients against the fractions of infected individuals.

### 5.3 Impact of varying *µ* and *µ*_0_ on listeriosis disease

To investigate the effect of *µ* and *µ*_0_ on the listeriosis spread, we simulate system (3) for different values of *µ* and *µ*_0_ against the infected and the aware individuals over the modelling time. Figure 4 depicts that an increase in the rate of implementation of awareness programs will lead to decrease in the number of infected individuals, and an increase in the number of aware individuals as they avoid being infected (see Figure 4). Also, a decrease in the diminution of awareness programs results in more human Listeria infection and a fewer number of aware individuals as shown in Figure 4(a) and 4(b) respectively. These results suggest that there is a need for awareness programs to educate the population about the risk of contracting listeriosis. This result is critical in the management and control of the disease.

**Figure 4:**
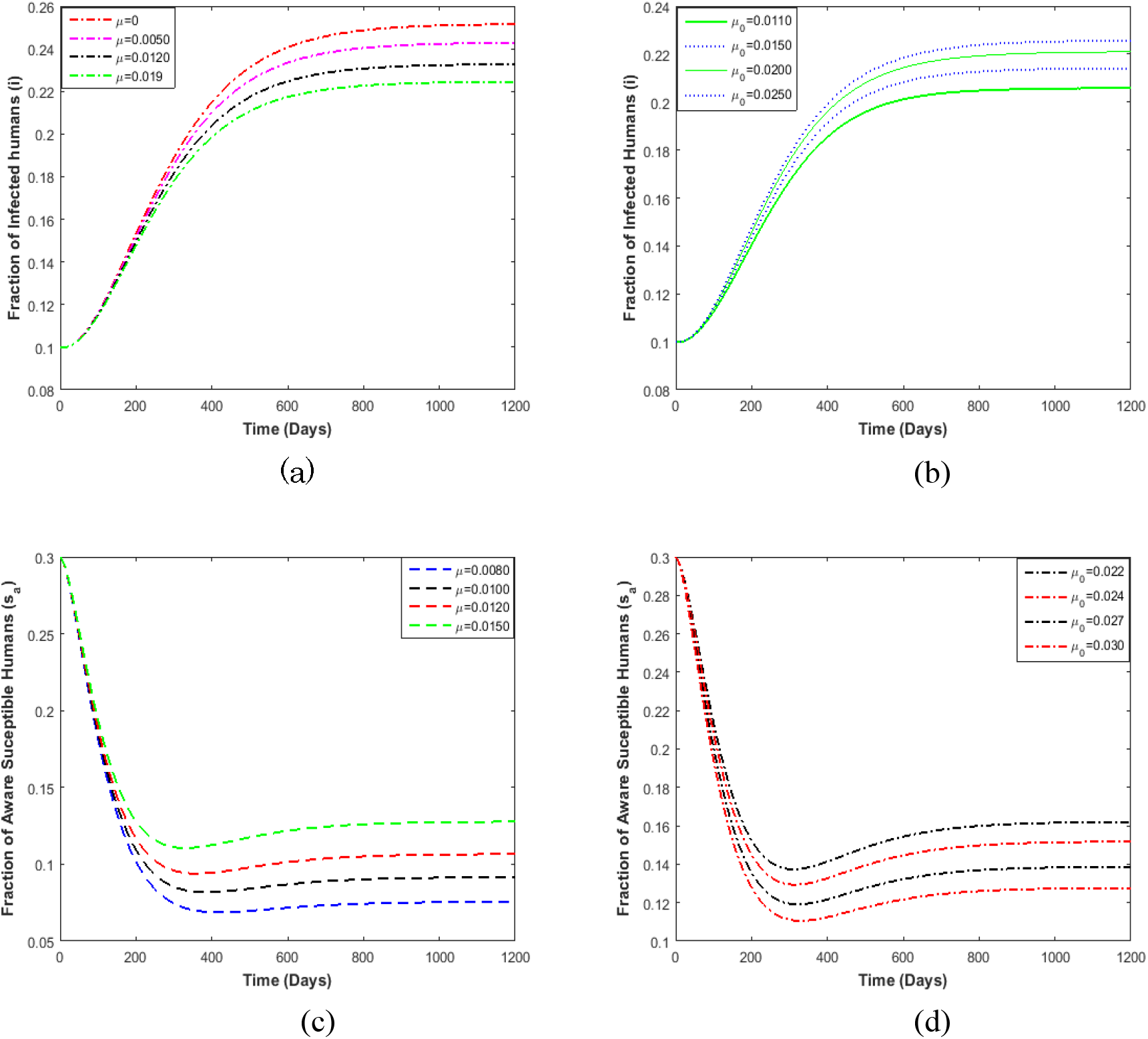
(a) and (b) Shows the effects of varying parameters; *µ* and *µ*_0_ on the fractions of *i* while Figure (a) and (b) shows the effects of varying parameters; *µ* and *µ*_0_ on *s*_*a*_-individuals respectively. The values of ℛ_*f*_ = 1.9143 with the rest parameter values as in Table 1.

### 5.4 Effects of *β*_3_ and *µ*_*f*_ on ℛ_*f*_

In this subsection, we investigate the effects of the rate of food contamination, *β*_3_, and the rate of food product removal, *µ*_*f*_ on the food contamination threshold ℛ_*f*_. This can be done by drawing a contour plot with *β*_3_ and *µ*_*f*_ on the axes. The contour plot shows an increase in *β*_3_, that is, the rate at which uncontaminated food is contaminated by contaminated food results in an increase in ℛ_*f*_. The result implies that more people will be infected with Listeria with increasing *β*_3_, while an increase in *µ*_*f*_ will cause fewer infections in humans, since the value of ℛ_*f*_ decreases. However, we emphasize that changes in the value of *β*_3_ impact ℛ_*f*_ less significant when compared to *µ*_*f*_.

### 5.5 Impact of varying *µ*_*f*_ and *ω* on *i* and *s*_*a*_ individuals

We observe in Figure 5 that increasing in *µ*_*f*_ decreases ℛ_*f*_. Thus, an increase in the values of *µ*_*f*_ results in a decrease in the number of infected humans. Figure 5, shows that a decrease in the rate at which the unaware humans, becomes aware of listeriosis (*ω*), results in more human infections. Similarly, we notice that increasing *ω* increases the number of aware individuals in the human populations as depicted in Figure 5.

**Figure 5:**
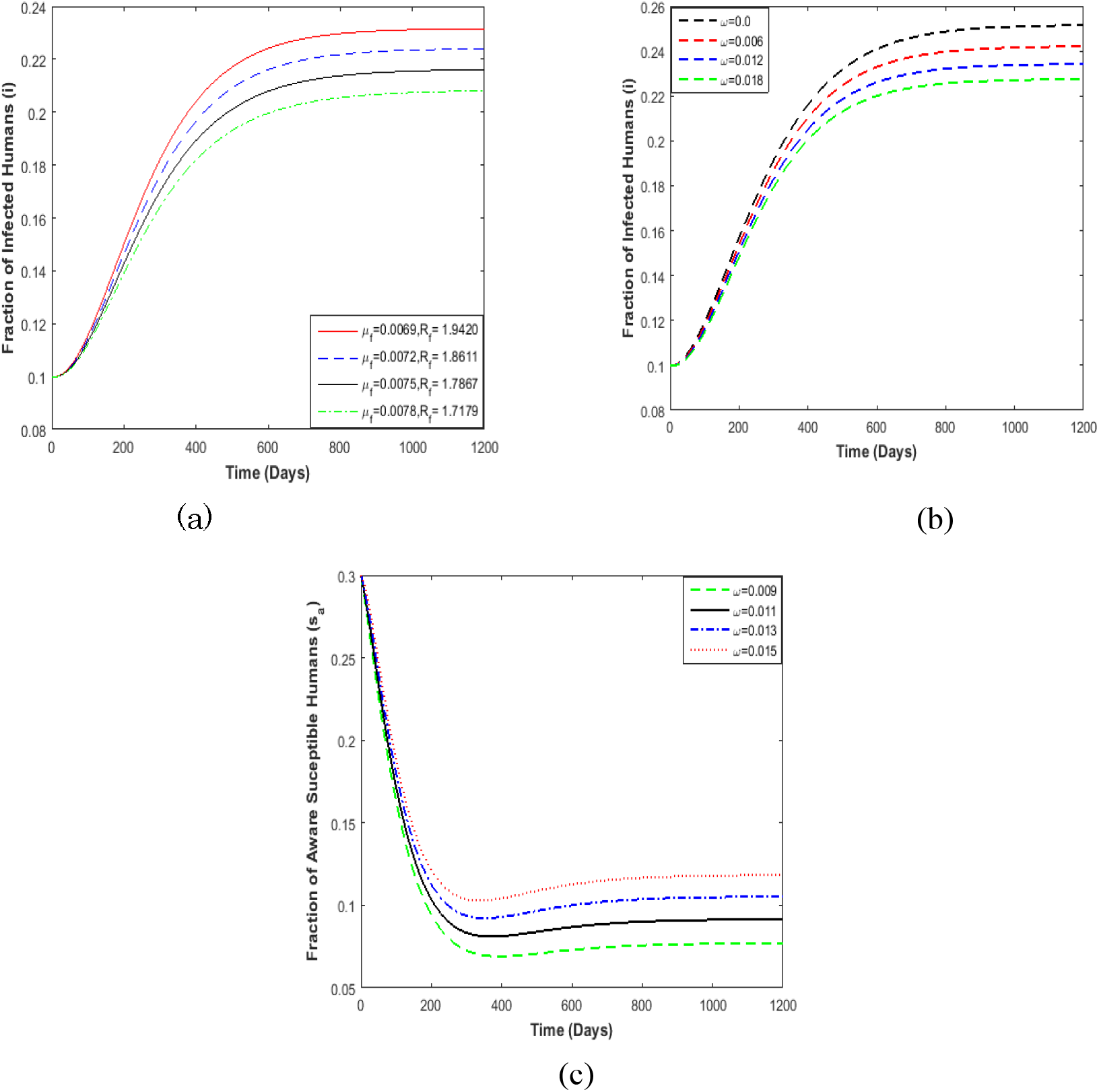
(a) Varying rate of food removal of food products (μ_*f*_) on the infected humans. Varying rate at which unaware susceptible becomes aware (*ω*) against. (b) Infected individual *i* and (c) aware individuals *s*_*a*_, with the rest parameters values as given in Table 1.

### 5.6 Varying *ε* on the fractions of infected and aware individuals

Figure 6 illustrates the effects of varying the efficacy parameter on the dynamics of the listeriosis. We see that an increase in the efficacy of awareness program over time reduces the number of individuals who become infected, see Figure 6. Also, an increase in *ε*, results in an increase in the number of aware individuals as depicted in Figure 6. This implies that increased implementation of awareness programs results in more humans becoming aware of the disease and take precautionary measures thereby avoiding being infected by Listeria.

**Figure 6:**
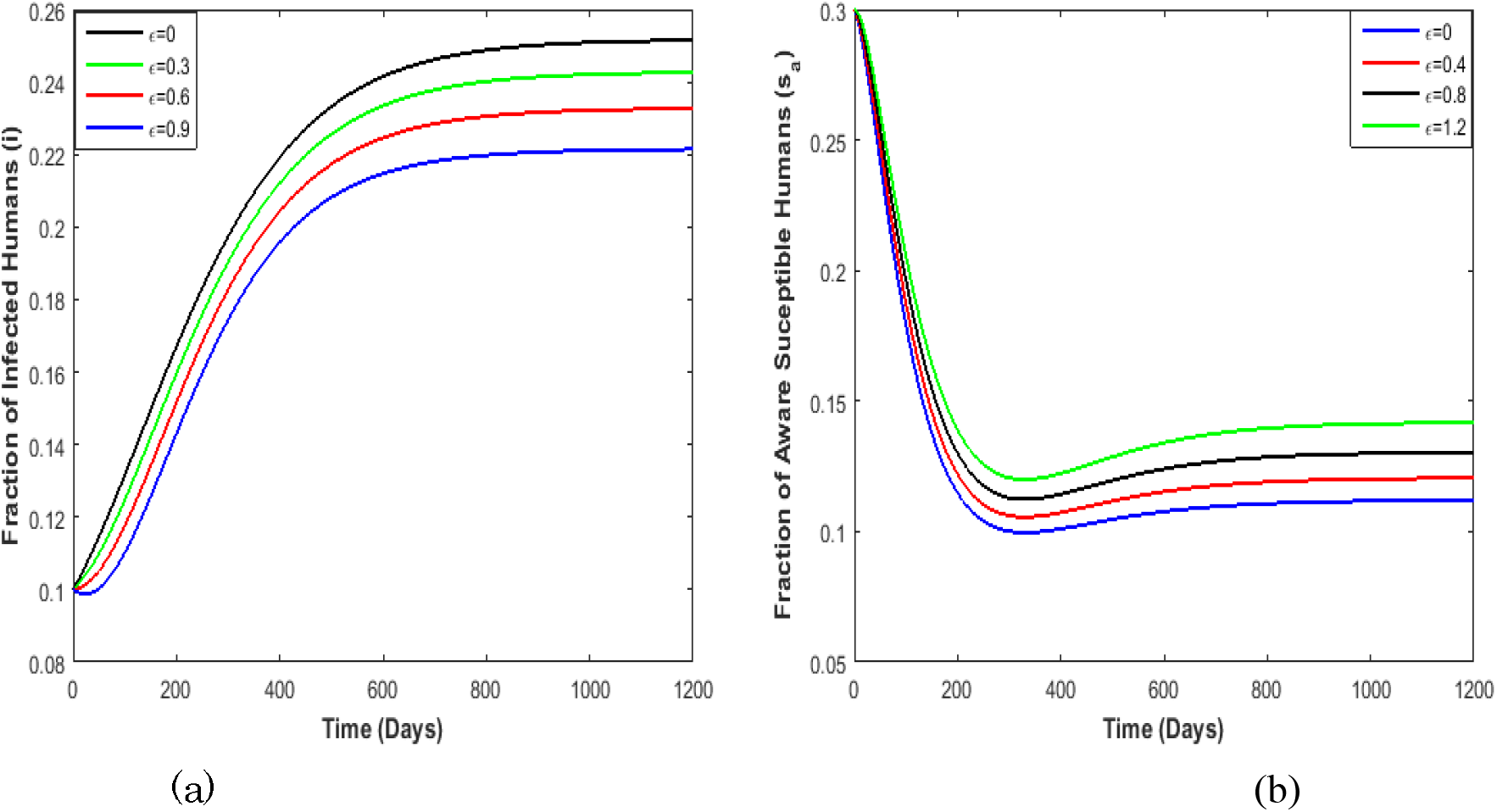
Varying efficacy of awareness programs parameter *ε* against (a) Fraction of infected humans *i*, and (b) aware susceptibles humans *s*_*a*_. The numerical value of ℛ_*f*_ is 1.9143. Note that *ϵ* represents *ε* in the legend.

## 6 Discussion and conclusion

The control of infectious diseases through awareness programs by media is important in reducing human mortality caused by foodborne diseases. In the present study, we used a mathematical model to study the impact of awareness programs on the spread or contracting listeriosis disease. It was assumed that aware susceptibles interact with the infectives due to ineffectiveness or waning of awareness programs over the modelling time. The equilibrium points of the model system (3) were determined and their local asymptotic stabilities were analysed using the food contamination threshold ℛ_*f*_. Further, the analytical findings in this manuscript were confirmed by some numerical simulations. Uncertainty sensitivity analysis was used to verify which parameters are sensitive to some of the state variables of interest. Results from the numerical simulations of most model sensitive parameters show that increase: in the efficacy of awareness programs *ε*, the removal rate of contaminated food products *µ*_*f*_, the rate at which the susceptible individuals becomes aware during disease spread will result in fewer listeriosis infections. The implications of these results are vital, in that, there is a need for constant implementation of awareness programs by the media at all times with very low waning rates. On the other hand, reducing the rate at which the rate of food contamination (*β*_3_) will prevent high food contamination, and therefore fewer human Listeria infections. The analytical and numerical results of our findings suggest the implementation of awareness programs that do not wane over time, if the humans practice proper hygiene, in other words, avoid contracting Listeria (bacteria) from the environment, and reduction in the consumption of contaminated food products results in fewer infections in the human populations.

The model presented in this paper has a number of shortcomings. The assumption that the human population is constant only works for epidemics with a short duration and listeriosis seems to have been in the human population for long. The model was also not fitted to epidemiological data. The model can be improved by considering a varying human population and age dependency in the infection. Despite these shortcomings, the results remain valid and implementable by policymakers or food safety risk management to contain or eradicate the disease during listeriosis outbreaks.

## Data Availability

No data utilized

## Author‘s contributions

CW Chukwu carried out the analytical, and numerical simulations. Both authors formulated and commented on the final version of this work.

## Acknowledgement‘s

The authors would like to thank the Faculty of Science at the University of Johannesburg for their support in the production of this manuscript. The authors are grateful to the anonymous reviewers for their lucrative suggestions and comments.

